# Unique prediction of developmental psychopathology from genetic and familial risk

**DOI:** 10.1101/2020.09.08.20186908

**Authors:** Robert Loughnan, Clare E. Palmer, Carolina Makowski, Wesley K. Thompson, Deanna M. Barch, Terry L. Jernigan, Anders M. Dale, Chun Chieh Fan

**Affiliations:** Department of Cognitive Science, University of California, San Diego, 9500 Gilman Drive, La Jolla, CA 92093, USA; Population Neuroscience and Genetics, University of California, San Diego, 9500 Gilman Drive, La Jolla, CA 92161, USA; Center for Human Development, University of California, San Diego, 9500 Gilman Drive, La Jolla, CA 92161, USA; Center for Multimodal Imaging and Genetics, University of California, San Diego School of Medicine, 9444 Medical Center Dr, La Jolla, CA 92037, USA; Herbert Wertheim School of Public Health and Human Longevity Science, University of California, San Diego, La Jolla, CA, USA; Psychological & Brain Sciences, Psychiatry and Radiology, Washington University in St. Louis, St. Louis USA; Department of Radiology, University of California, San Diego School of Medicine, 9500 Gilman Drive, La Jolla, CA 92037, USA; Department of Neuroscience, University of California, San Diego School of Medicine, 9500 Gilman Drive, La Jolla, CA 92037, USA; Department of Psychiatry, University of California, San Diego School of Medicine, 9500 Gilman Drive, La Jolla, CA 92037, USA

**Author notes:** Corresponding authors: Robert Loughnan, Center for Human Development, La Jolla, CA 92093, Chun Chieh Fan, Center for Human Development, 9500 Gilman Drive, La Jolla, CA 92093. These authors contributed equally to this work (Robert Loughnan, Clare E. Palmer).

## Abstract

**Background:** Early detection is critical for easing the rising burden of psychiatric disorders. However, the specificity of psychopathological measurements and genetic predictors is unclear among youth.

**Methods:** We measured associations between genetic risk for psychopathology (polygenic risk scores (PRS) and family history (FH) measures) and a wide range of behavioral measures in a large sample (n=5204) of early adolescent participants (9-11 years) from the Adolescent Brain and Cognitive Development (ABCD) Study^SM^. Associations were measured both with and without taking into consideration shared variance across measures of genetic risk.

**Results:** Polygenic risk for Attention Deficit Hyperactivity Disorder (ADHD) and depression (DEP) shared many significant associations with externalizing, internalizing and psychosis-related behaviors. However, when accounting for all measures of genetic and familial risk these two PRS also showed clear, unique patterns of association: the DEP PRS showed significantly stronger associations with somatic complaints and depression symptoms; whereas the ADHD PRS showed stronger associations with ADHD symptoms, impulsivity and prodromal psychosis. The Schizophrenia PRS showed a unique negative association with performance on cognitive tasks measuring fluid abilities, such as working memory and executive function, that was not accounted for by other measures of genetic risk. FH accounted for unique variability in behavior above and beyond PRS and vice versa with FH measures explaining a greater proportion of unique variability compared to the PRS.

**Conclusion:** Our results indicate that, among youth, many behaviors show shared genetic influences; however, there is also specificity in the profile of emerging psychopathologies for individuals with high genetic risk for particular disorders. This may be useful for quantifying early, differential risk for psychopathology in development.

**Funding:** The ABCD Study is supported by the National Institutes of Health and additional federal partners under award numbers U01DA041022, U01DA041028, U01DA041048, U01DA041089, U01DA041106, U01DA041117, U01DA041120, U01DA041134, U01DA041148, U01DA041156, U01DA041174, U24DA041123, U24DA041147, U01DA041093, and U01DA041025. A full list of supporters is available at https://abcdstudy.org/federal-partners.html. R.L was supported by Kavli Innovative Research Grant under award number 2019-1624. C.F. was supported by grant R01MH122688 and RF1MH120025 funded by the National Institute for Mental Health (NIMH).

## INTRODUCTION

Psychiatric disorders place a huge burden on those affected, their families and society. Identifying risk for psychopathology in developmental samples may offer an opportunity for early detection and intervention. Nearly all psychiatric disorders have a heritable component, with twin heritability estimates ranging from 33-84% across affective, psychotic and developmental disorders^1^. Lifetime prevalence rates of several disorders are higher among first degree biological relatives of individuals with a diagnosis compared to families of individuals with no diagnosis^2^. Therefore, estimating genetic liability for psychiatric disorders presents one avenue for identifying at risk individuals and probing differential and transdiagnostic risk factors. Here we sought to determine: 1) if increased genetic risk within a large, typically developing and demographically diverse developmental sample would be associated with symptoms of psychopathology, related individual difference factors, and cognitive function; and, 2) whether there was any evidence for specificity in the behavioral measures predicted by different genetic markers.

Large-sample analyses of results from genome-wide association studies (GWAS) have revealed the highly polygenic architecture of complex behavioral phenotypes, with many variants in the genome additively accounting for substantial heritability, but individually exerting only very small effects. Models using effect sizes at single nucleotide polymorphisms (SNPs) estimated from large-scale independent GWAS, can be used to compute polygenic risk scores (PRS), which are aggregate scores of an individual’s genetic risk for a trait. Notably recent powerful, cross-disorder meta-analyses ^3,4^ reveal high genetic correlation and widespread pleiotropy across psychiatric disorders, consistent with overlapping genetic architecture. Indeed, polygenic risk for depression has been shown to positively associate with childhood psychopathology across behavioral domains^5^.

Family history (FH) is a clinically used factor for predicting psychiatric risk^6^, yet there has been a lack of direct comparisons of associations between PRS and FH of psychopathology in childhood and adolescence. SNP heritabilities (h_SNP_^2^) based on effects across the genome are lower than twin heritabilities, which suggests there are genetic factors driving psychiatric phenotypes that are not fully captured with common variants at current GWAS sample sizes. Indeed, FH likely reflects a complex combination of genetic and environmental factors. Due to the differential information that PRS and FH measures may provide, it is important to determine whether they explain independent or overlapping variance in developmental psychopathology and cognition. For example, in large cohorts, both family history of schizophrenia ^7^ and polygenic risk for schizophrenia ^8^ have been shown to associate with developmental psychopathology; however, the unique contribution of polygenic risk above and beyond FH is unclear.

For this study we used behavioral and genetic data from 9-11 year-old children from the Adolescent Brain and Cognitive Development (ABCD) Study^SM^. We generated five PRS that were trained on large independent datasets. We used these PRS and measures of FH of psychopathology both independently and within the same models to predict a large array of both caregiver and youth-reported phenotypes thought to reflect behavioral risk for developing psychiatric disorders. These measures included both dimensional and diagnostic assessments of psychopathology, individual difference measures of impulsivity and behavioral approach and inhibition, prodromal psychosis and behaviors associated with mania and prosocial behavior. We additionally measured associations with cognitive measures from the NIH Toolbox given documented associations and genetic overlap between cognitive impairment and schizophrenia and bipolar disorder^9,10^. Using this approach, we aimed to uncover variability across early signs of psychopathology across domains that is uniquely associated with each genetic/familial predictor. This research is an essential first step in this large longitudinal study to determine whether we can identify early signs of specificity in genetic-behavior associations in development, which can then be tracked to determine their potential predictive power for future diagnoses.

## METHODS & MATERIALS

### Sample

The ABCD study is a longitudinal study across 21 data acquisition sites in the United States following 11,880 children starting at 9-11 years. This paper uses baseline data from the NIMH Data Archive ABCD Collection Release 2.0.1 (DOI: 10.15154/1504041). The ABCD cohort was recruited to ensure the sample was as close to nationally representative as possible, therefore the cohort includes individuals across different racial and ethnic backgrounds and socioeconomic groups ^11^. There is an embedded twin cohort and many siblings. As the chosen PRS were all trained on European individuals, the main associations in this study were conducted in a predominantly European ancestry sample (n=5204). Supplementary analyses were conducted in those with non-European ancestry (n=3964) and the full sample (n=9168). Table 1 outlines the demographics of the European sample used in the main analysis.

**Table 1.** Sociodemographic breakdown for the European ancestry sample analyzed in this study. All main analyses were conducted with a European ancestry only sample. Compared to the full ABCD sample had higher proportions of individuals from households with higher income and a higher parental education level and self-identifying as White. Age, sex, household income, parental education, data collection site and the top 10 genetic principal components were controlled for in the main analyses. Supplementary analyses were conducted without controlling for SES and in the Full sample and Non-European ancestry sample.

|  | **European Ancestry Sample** |
| --- | --- |
| **Total N** | 5,204 |
| **Singletons n** | 4,418 |
| **Age - months (%)** | 119.18 (7.48) |
| **Sex, n (%)** |  |
| M | 2744 (52.7) |
| **Parental Education, n (%)** |  |
| < HS Diploma | 22 (0.4) |
| HS Diploma/GED | 149 (2.9) |
| Some College | 963 (18.5) |
| Bachelor | 1650 (31.7) |
| Post Graduate Degree | 2420 (46.5) |
| **Household Income, n (%)** |  |
| [<50K] | 642 (12.3) |
| [>=50K & <100K] | 1590 (30.6) |
| [>=100K] | 2972 (57.1) |
| **Race Ethnicity, n(%)** |  |
| White | 4934 (94.9) |
| Black | 1 (0.0) |
| Hispanic | 137 (2.6) |
| Asian | 0 (0.0) |
| Other | 126 (2.4) |

### ABCD Baseline Mental Health Battery

The Mental Health Battery in ABCD is an extensive battery of questionnaires and semi-structured interviews assessing diagnostic and dimensional measures of psychopathology and individual difference factors. Both youth and their caregivers provided responses at baseline using divergent and overlapping measures. Motivation behind selecting these assessments is outlined here: ^12^. Supplementary Table 1 lists variables used from the ABCD public release.

### Diagnostic Assessments

#### Kiddie Schedule for Affective Disorders and Schizophrenia (KSADS)

Participants completed a semi-structured, self-administered, computerized version of the validated and reliable KSADS-5^13^. Research Assistants had extensive training to support youth completing this assessment. Caregivers and youth completed modules on depression, bipolar disorder, generalized anxiety disorder, social anxiety disorder, suicidality and sleep. Only caregivers completed psychosis, obsessive-compulsive disorder (OCD), ADHD, oppositional defiant disorder (ODD), conduct disorder (CD), panic disorder and eating disorders modules. Symptom scores were the sum of lifetime symptoms endorsed in each module and were scored and analyzed separately for each respondent. The total symptom score was a sum across modules.

### Dimensional Assessments

#### Child Behavior Checklist (CBCL)

Caregiver-reported CBCL^14^ has eight syndrome scales: anxious/depressed, withdrawn/depressed, somatic complaints, social problems, thought problems, attention problems, rule breaking behavior and aggressive behavior, and a total problems score.

#### General behavior inventory

Caregiver-report ten-item Mania Scale^15^ derived from the 73-item General Behavior Inventory (PGBI) for Children and Adolescents^16^.

#### Prosocial Behavior Survey

Caregivers and youth were asked three questions about how helpful and considerate the youth was in general, with summed scores for both caregiver and youth.

#### Prodromal Questionnaire Brief (PQ-B)

Youth-report measure, modified for use in children in our age range, consisting of a 21-item scale assessing subclinical manifestations of psychosis^17–19^. The prodromal psychosis severity score is the sum of the number of symptoms endorsed weighted by how distressing the symptoms were.

#### UPPS-P for children short scale

Youth-report impulsive behavior scale, which includes five sub-scales that measure four factors of impulsivity: positive and negative urgency, lack of perseverance, premeditation, and sensation seeking^20^.

#### Behavioral inhibition and behavioral activation (BISBAS scale)

Youth-report measure of approach and avoidance behaviors ^21,22^ that produces scores for drive, fun seeking, reward responsiveness, and behavioral inhibition.

#### NIH Toolbox Cognition Battery®

Widely used battery of cognitive tests that measures a range of different cognitive domains^23–25^. We analyzed the uncorrected composite scores broadly measuring fluid and crystallized intelligence that are generated from the NIH Toolbox® and have been validated against gold-standard measures ^26–28^. The fluid composite score includes performance on the flanker task, picture sequence memory task, list sorting memory task, pattern comparison processing speed and dimensional change card sort task. The crystallized composite score includes performance on the oral reading recognition task and picture vocabulary task.

### Genetic & Familial Measures

#### Genetic Data Preprocessing

*DNA samples were collected at baseline visits using either blood or saliva samples. Genotyping was performed on 646,247 genetic variants using the Affymetrix smokescreen array*^29^. *Imputation of variants was performed using the Michigan Imputation Server*^30^ *resulting in a total of 1,427,972 variants on 10,659 individuals passing quality control measures. For further details on imputation, quality control measures and ancestry estimation of genetic data please see supplementary materials*.

#### Polygenic Risk Scores (PRS)

PRS were estimated from summary statistics for ADHD^31^, Autism Spectrum Disorder (ASD)^32^, Bipolar Disorder (BPD)^33^, Schizophrenia (SCZ)^34^ and Depression (DEP)^35^ from the Psychiatric Genetics Consortium (https://www.med.unc.edu/pgc/results-and-downloads). Additional details of preprocessing genetic data and PRS estimation are in supplementary materials.

#### Family History Assessment

*Caregivers were given a questionnaire asking about family history (FH) of 10 behaviors associated with psychopathology: alcohol use; drug use; depression; mania; psychosis; conduct problems; nerves; seen a therapist; hospitalized for a mental health problem; and, suicide. For each question the caregivers were asked specifically if any blood relative had experienced any of the described behaviors (more detail in Supplementary Table 2). Importantly, these variables do not indicate clinical diagnoses associated with these behaviors*.

### Statistical Analysis

Generalized Linear Models (GLMs) were fit to measure the association between i) each of the 41 behavioral phenotypes and ii) FH and PRS. Univariate models included one independent variable of interest (PRS or FH) in each model (i.e. behavior ∼ PRS_i_ + covariates or behavior ∼ FH_i_ + covariates). Multivariable models included all PRS and FH measures in the same model (i.e. behavior ∼ PRS_1_ +PRS_2_ … +FH_1_ + FH_2_ … + covariates). Each behavioral phenotype was modelled separately with the same set of predictors and covariates. Fixed nuisance covariates included age, sex, top 10 genetic principal components, household income, highest parental education and data collection site. ΔR^2^ was reported as change in R^2^ from a reduced model (covariates only) to a full model (including the predictor of interest)^36,37^. Supplementary analyses were conducted without controlling for household income and parental education to understand the impact of socioeconomic status (SES). Family relatedness was controlled for using a random subset of the sample that only included singletons. Significant associations were determined using a false discovery rate (FDR) and reported p-values are FDR adjusted (p-adj). See supplement for additional analysis details. Additional models were implemented to measure pairwise spearman correlations across all of the DVs and IVs in the European ancestry sample after residualizing for the covariates of no interest (Supplementary Figures 2&3). Behavioral measures were categorized by behavioral domain (see Supplementary Table 3) in order to determine whether associations with each genetic predictor were enriched for measures within domains.

## RESULTS

### Unique behavioral associations with PRS across domains

In the univariate models (measuring the association between each PRS and each behavioral variable in separate models), controlling for SES, the ADHD and DEP PRS showed the largest and greatest number of associations across internalizing, externalizing and psychosis-related measures (Figure 1, left panel). The ADHD PRS significantly associated with CBCL rule-breaking (ΔR^2^=0.0071, p-adj=5.7⨯10^−6^), inattentive (ΔR^2^=0.0064, p-adj=4.6⨯10^−8^) and aggressive (ΔR^2^=0.0031, p-adj=5.2⨯10^−4^) behaviors, prodromal psychosis severity (ΔR^2^=0.0062, p-adj=2.4⨯10^−5^), and caregiver reported KSADS oppositional/conduct disorder (ΔR^2^=0.0042, p-adj=7.7×10^−5^) and ADHD (ΔR^2^=0.0030, p-adj=9.2×10^−4^) symptoms, followed multiple youth and caregiver reported measures of impulsivity, depression and suicidality symptoms, bipolar and psychosis related measures and developmental social problems. The DEP PRS showed largest significant associations with CBCL somatic complaints (ΔR^2^=0.0054, p- adj=1.9⨯10^−6^), KSADS symptoms of oppositional/conduct disorder (ΔR^2^=0.0039, p- adj=1.6×10^−4^) and CBCL anxious/depressive (ΔR^2^=0.0031, p-adj=2.5×10^−4^), aggressive (ΔR^2^=0.0031, p-adj=5.1×10^−4^), and rule-breaking (ΔR^2^=0.0029, p-adj=4.8×10^−6^) behaviors. These were followed by caregiver reported KSADS symptoms of suicidality (ΔR^2^=0.0027, p-adj=1.8⨯10^−3^), bipolar disorder (ΔR^2^=0.0027, p-adj=2.0⨯10^−3^) and anxiety (ΔR^2^=0.0020, p-adj=8.4⨯10^−3^) and youth reported KSADS depression symptoms (ΔR^2^=0.0027, p-adj=2.2⨯10^−3^), as well as other measures of negative urgency, developmental social problems, behavioral inhibition and bipolar and psychosis related behaviors. Both the ADHD and DEP PRS were also associated with the CBCL Total Problems and KSADS Total Symptoms scores.

**Figure 1.**
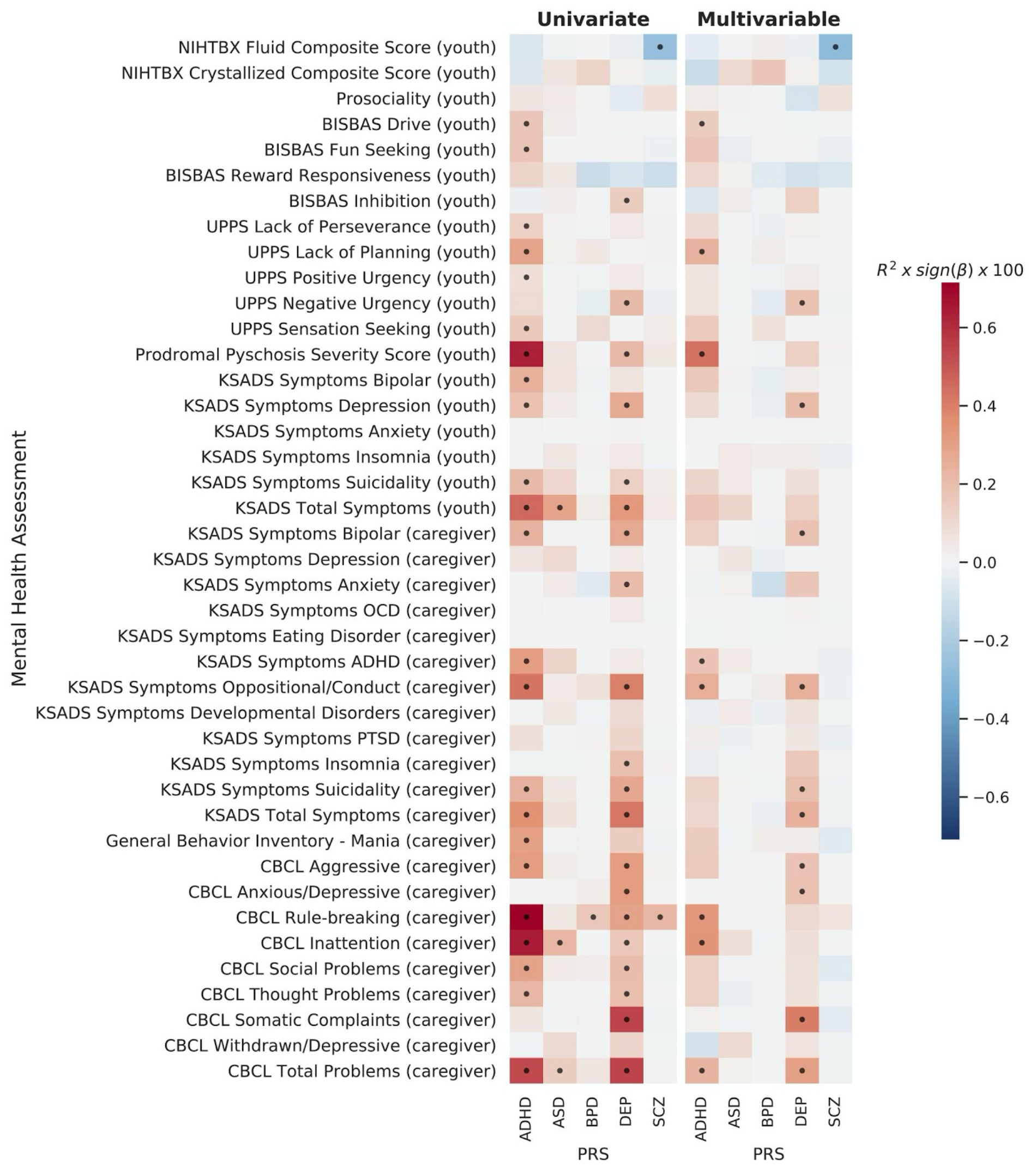
Univariate (left) and multivariable (right) associations for each behavioral phenotype predicted by the PRS. Effect sizes for each association are displayed as the partial variance explained, R2, (as a percentage) multiplied by the sign of the beta estimate to indicate the magnitude and sign of the association (red=positive, blue=negative). Each row represents a model with the dependent variable along the y-axis and each PRS on the x-axis. In the univariate models (left) only a single genetic predictor was included in each model (each cell = 1 model) – i.e. behavior ∼ PRS + covariates. In the multivariable models (right) all genetic/familial predictors were included in each model including all PRS and FH measures (each row = 1 model) – i.e. behavior ∼ PRS_1_ +PRS_2_ … +FH_1_ + FH_2_ … + covariates. Along the x-axis from left to right: the five PRS measured (Attention Deficit Hyperactivity Disorder (ADHD), Autism Spectrum Disorder (ASD), Bipolar Disorder (BPD), Depression (DEP), Schizophrenia (SCZ)). All models controlled for covariates of age, sex, the top 10 Principal Components of the genetic data, household income, highest parental education and data collection site. Effects represent the median across 100 subsamples of singletons to control for family relatedness. Dots indicate FDR significant associations.

The BPD and SCZ PRS were not significantly associated with any bipolar or psychosis-related measures; however, they did significantly associate with CBCL rule-breaking with a smaller effect size compared to ADHD and DEP (BPD: ΔR^2^=0.0016, p- adj=4.3⨯10^−2^; SCZ: ΔR^2^=0.0022, p-adj=1.5×10^−2^). In addition, the SCZ PRS negatively associated with the fluid composite score from the NIH Toolbox® (ΔR^2^=0.0027, p- adj=2.2×10^−3^). The ASD PRS was associated with CBCL inattention (ΔR^2^=0.0022, p- adj=1.9⨯10^−2^), youth reported KSADS total symptoms scale (ΔR^2^=0.0028, p-adj=1.4⨯10^−2^) and the CBCL total symptoms scale (ΔR^2^=0.0014, p-adj=3.1⨯10^−2^).

Multivariable models determined the specificity of these associations by covarying for all PRS and FH predictors simultaneously. In these models, PRS associations were attenuated and showed greater specificity for the ADHD and DEP PRS (Figure 1, right panel). The ADHD PRS predicted unique variance across externalizing and psychosis-related measures not predicted by other measures of genetic risk (PRS or FH), whereas the DEP PRS predicted unique variance across a different set of internalizing and externalizing behaviors. Additionally, in the multivariable models, only the ADHD PRS was significantly associated with CBCL rule-breaking. The ASD PRS also no longer showed any significant associations. The SCZ PRS association with the fluid composite score from the NIH Toolbox® remained significant when controlling for all other measures of genetic risk (ΔR^2^=0.0029, p-adj=7.7⨯10^−3^).

When not controlling for SES, behavioral associations were slightly larger and the overall pattern of associations was similar (Supplementary Figure 4,5). However, there were additional significant associations with cognitive measures for the ADHD and BPD PRS. The SCZ association with the fluid composite score was the only association with cognitive performance that was robust to controlling for SES.

We categorized each behavior into a domain based on the construct measured in order to highlight the different types of behavioral measures predicted by each PRS (Figure **2**; variables in each domain are highlighted in Supplementary Table 1). Across both the univariate and multivariable models, the largest associations with the ADHD PRS were with externalizing and psychosis-related measures; whereas for the DEP PRS, associations encompassed a mix of internalizing and externalizing measures. In the multivariable models, the specificity in the unique pattern of behaviors predicted by these PRS across domains was clarified due to the removal of any shared variance across the genetic predictors.

**Figure 2.**
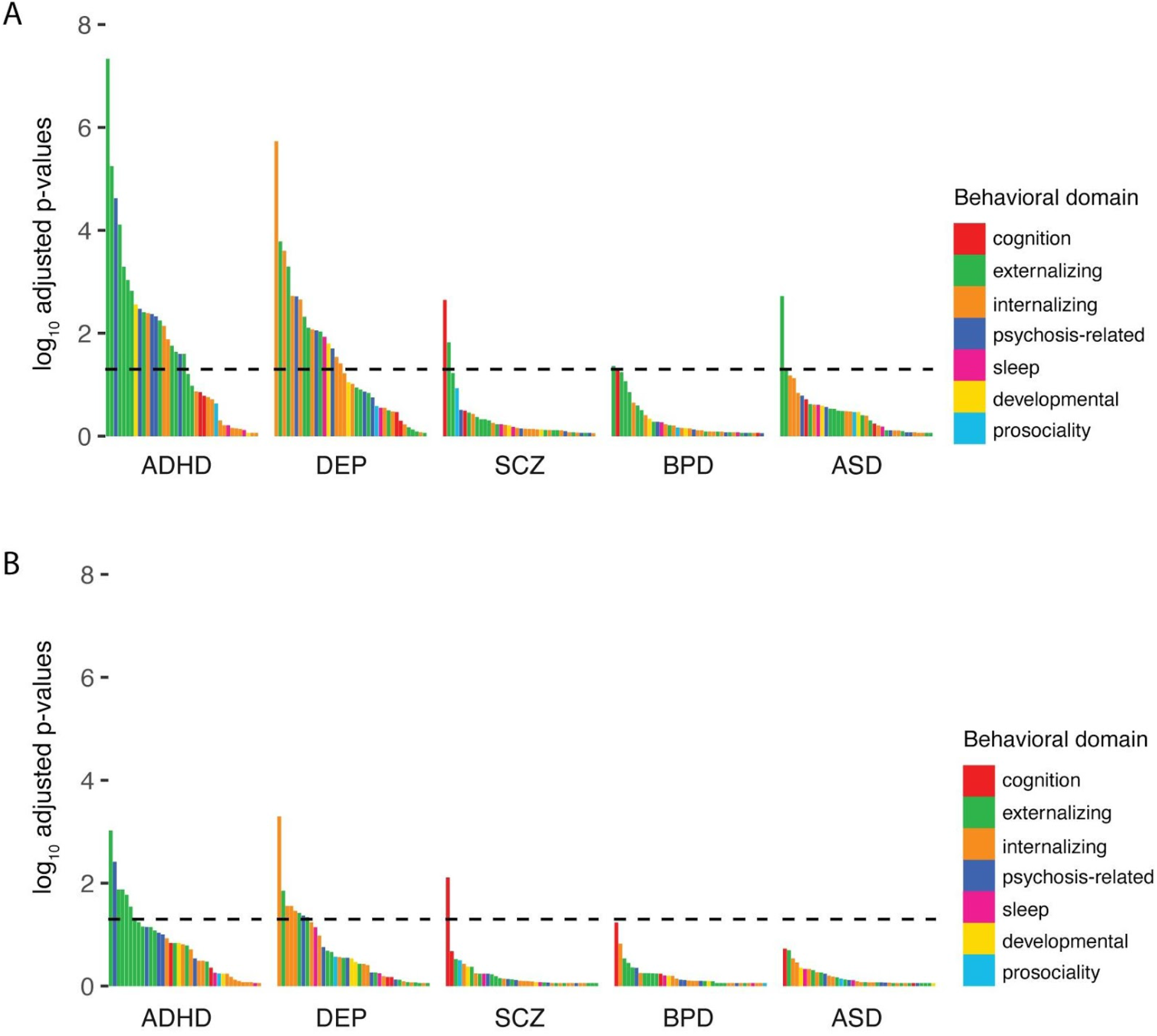
Enrichment of PRS associations across different behavioral domains. FDR adjusted p-values (logged) for all of the associations shown in Figure 1 for both the univariate (A) and multivariable models (B). Each association is colored based on the behavioral domain the dependent variable was categorized within (see supplementary table 3). Associations are ordered with the most significant effect to the far left. The horizontal line represents the FDR adjusted significance threshold (p=0.05). All models controlled for covariates of age, sex, the top 10 Principal Components of the genetic data, household income, highest parental education and data collection site. Effects represent the median across 100 subsamples of singletons to control for family relatedness.

#### Unique behavioral associations with FH across domains

Behavioral associations with FH measures were larger than with PRS (Figure 3, left panel) in the univariate models. Given the large number of overlapping univariate associations, we will focus on the associations from the multivariable models (i.e. controlling for all other FH and PRS predictors). In the multivariable models, FH of conduct problems, depression and anxiety/stress showed the largest effects with some specificity across the behavioral measures (Figure 3, right panel). FH of conduct problems significantly associated with the CBCL subscales particularly with rule-breaking (ΔR^2^=0.0081, p-adj=1.8⨯10^−5^), as well as KSADS symptoms related to both externalizing and internalizing disorders (ΔR^2^range=0.0023-0.0072), and mania (ΔR^2^=0.0057, p-adj=8.0×10^−3^). FH of depression significantly associated with the total problems scales from the CBCL (R2=0.0046, p-adj=4.5⨯10^−4^) and KSADS (ΔR^2^=0.0043, p-adj=5.8⨯10^−4^), as well as internalizing and externalizing measures across the KSADS and CBCL (ΔR^2^range=0.0018-0.0040). This pattern was similar to DEP PRS, however, unlike the DEP PRS, FH of depression only associated with caregiver-reported measures in the multivariable models. FH of anxiety/stress showed several associations across domains with the largest effects for caregiver-reported KSADS anxiety symptoms (ΔR^2^=0.0081, p-adj=5.8×10^−7^) and the CBCL anxious/depressive subscale (ΔR^2^=0.0071, p-adj=5.8⨯10^−7^).

**Figure 3.**
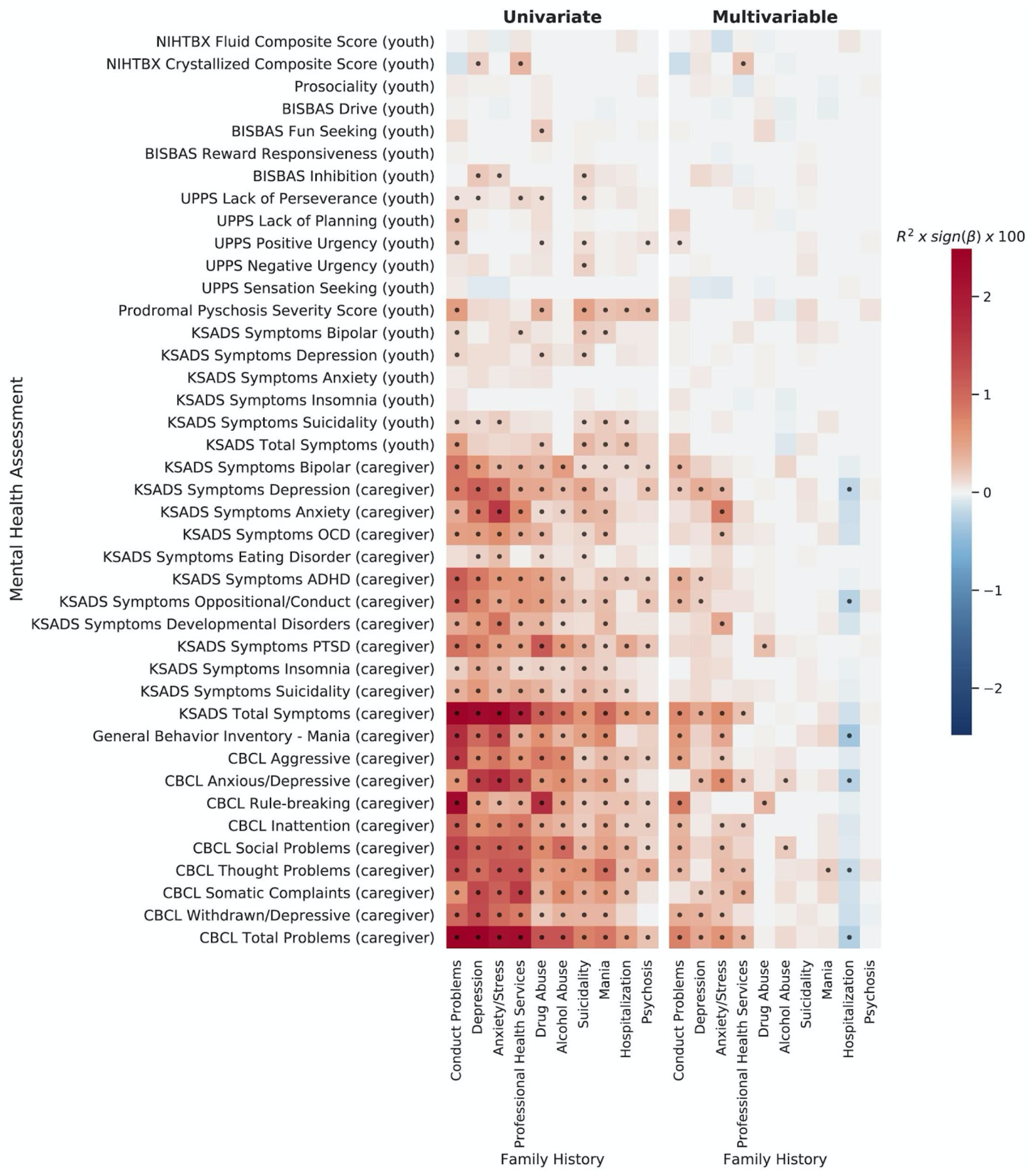
Univariate (left) and multivariable (right) associations for each behavioral phenotype predicted by FH. Effect sizes for each association are displayed as the partial variance explained, R2, (as a percentage) multiplied by the sign of the beta estimate to indicate the magnitude and sign of the association (red=positive, blue=negative). Each row represents a model with the dependent variable along the y-axis and each FH measure on the x-axis. In the univariate models (left) only a single familial predictor was included in each model (each cell = 1 model) – i.e. behavior ∼ FH_i_ + covariates. In the multivariable models (right) all genetic/familial predictors were included in each model including all PRS and FH measures (each row = 1 model) – i.e. behavior ∼ PRS_1_ +PRS_2_ … +FH_1_ + FH_2_ … + covariates. All models controlled for covariates of age, sex, the top 10 Principal Components of the genetic data, household income, highest parental education and data collection site. Effects represent the median across 100 subsamples of singletons to control for family relatedness. Dots indicate FDR significant associations.

FH of use of professional health services was most strongly associated with CBCL somatic complaints (ΔR^2^=0.0040, p-adj=5.6×10^−4^), thought problems (ΔR^2^=0.0028, p- adj=7.0⨯10^−3^) and the total problem score (ΔR^2^=0.0036, p-adj=2.2⨯10^−3^), and also showed a positive association with the crystallized composite score (ΔR^2^=0.0027, p- adj=1.0⨯10^−2^). Interestingly, when controlling for all other measures of genetic risk, FH of drug and alcohol abuse associated with differential behaviors, with FH of drug abuse explaining unique variance in CBCL rule-breaking (ΔR^2^=0.0034, p-adj=1.2⨯10^−2^) and KSADS PTSD symptoms (ΔR^2^=0.0029, p-adj=7.6⨯10^−3^), and FH of alcohol abuse explaining unique variance in CBCL social problems (ΔR^2^=0.0020, p-adj=4.9×10^−2^) and anxious/depressive behaviors (ΔR^2^=0.0017, p-adj=3.4×10^−2^). FH of hospitalization showed several negative associations with caregiver-reported internalizing behaviors, which were positive in the univariate models. This sign flip of effects may be due to collinearity across the genetic risk measures (Supplementary Figure 3) when used in a single model.

Figure **4** displays the enrichment of FH associations by behavioral domain. For the univariate models, the FH measures associated with behaviors across several domains. These patterns became more specific towards particular domains in the multivariable models (controlling for other FH measures and the PRS). For example, FH of depression or anxiety/stress were significantly associated with internalizing behaviors, whereas FH of conduct disorder was significantly associated with externalizing behaviors.

**Figure 4.**
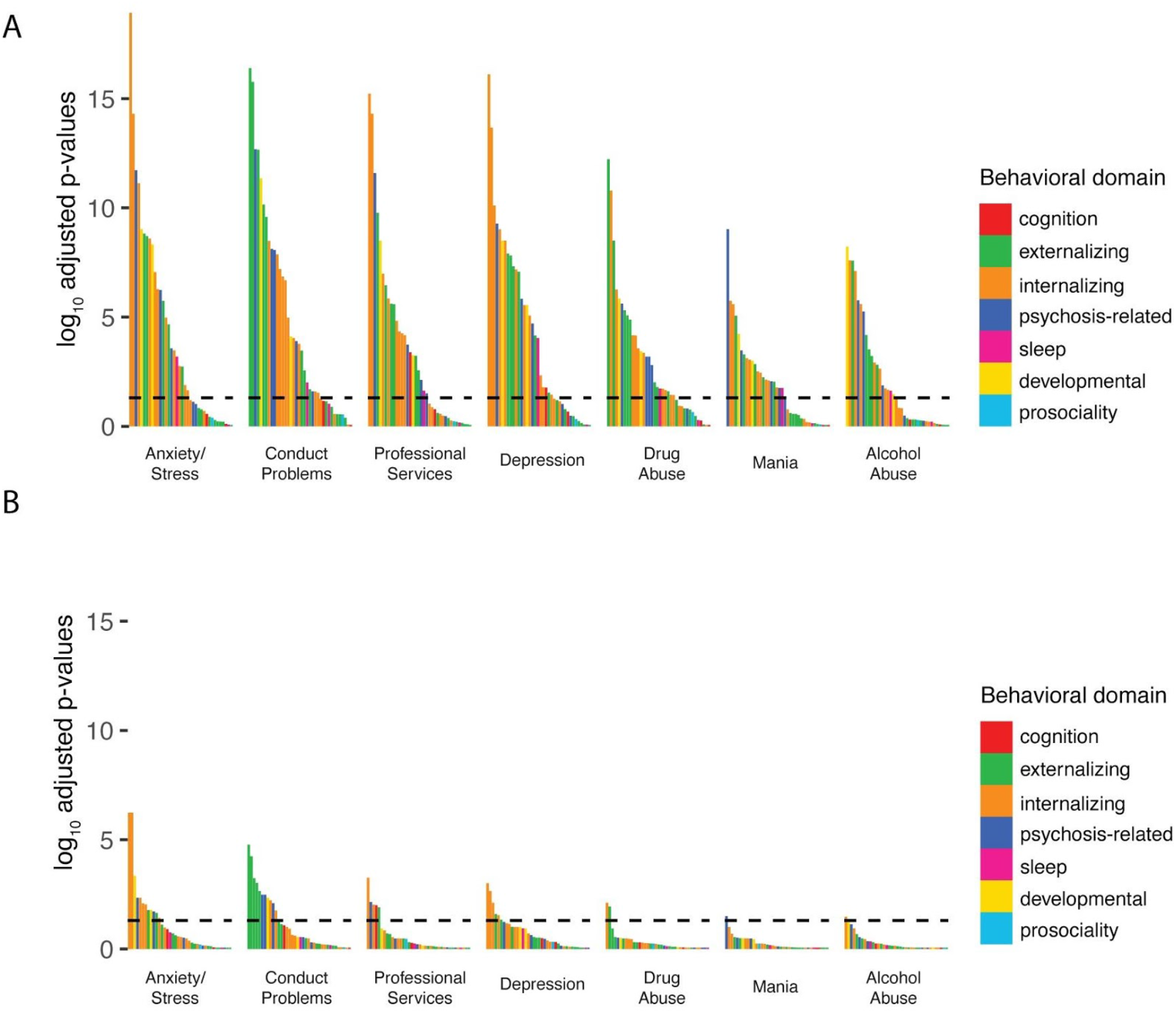
Enrichment of FH associations across different behavioral domains. FDR adjusted p-values (logged) for all of the associations shown in Figure 3 for both the univariate (A) and multivariable models (B). Each association is colored based on the behavioral domain the dependent variable was categorized within (see supplementary table 3). Associations are ordered with the most significant effect to the far left. The horizontal line represents the FDR adjusted significance threshold (p=0.05). All models controlled for covariates of age, sex, the top 10 Principal Components of the genetic data, household income, highest parental education and data collection site. Effects represent the median across 100 subsamples of singletons to control for family relatedness.

Finally, we quantified the variance in each behavior predicted by the set of PRS and set of FH measures when controlling for the other set of genetic predictors. Supplementary Table 3 shows that, in all cases, each set independently predicted unique variance over and above the other set of genetic predictors. The maximum variance explained by the FH and PRS measures combined was ΔR^2^=0.062 of CBCL Total Problems scale, of which ΔR^2^=0.053 was uniquely predicted by FH and ΔR^2^=0.0061 was uniquely predicted by PRS. The maximum unique variance explained collectively by PRS was ΔR^2^=0.0071 of the variability in oppositional/conduct disorder symptoms. These results further demonstrate that PRS and FH predict unique, non-overlapping variance across different domains of behavior in youth with PRS predicting a smaller proportion of variability than FH.

## DISCUSSION

Polygenic risk and FH of psychopathology predict both overlapping and unique variability in behavior across domains in 9-11 year old youth. Several externalizing and internalizing behaviors were associated with multiple measures of genetic risk highlighting shared genetic influences underlying variability in developmental psychopathology. However, when controlling for shared variance across PRS and FH measures, polygenic risk for ADHD and depression predicted unique variance across differential externalizing, internalizing and psychosis-related behaviors with the strongest associations for ADHD and depression symptomatology. Moreover, the SCZ PRS specifically and uniquely predicted variability in cognitive performance. This highlights that these PRS are signaling differential behavior related to specific disorders. FH of psychopathology explained additional unique variance in behavior, independent of the PRS, indicating additional genetic and environmental influences on behavior and recapitulating results in adults demonstrating the complementary information provided by PRS and FH^38,39^. Using the combined information across these genetic and familial measures and the dense behavioral phenotyping in the ABCD study, we have identified several, specific patterns of behavior associated with genetic risk for psychopathology that may be useful for quantifying early risk across different disorders during development.

Of the PRS analyzed, the ADHD and DEP PRS showed univariate associations across largely overlapping behavioral measures. In particular, both PRS predicted variability in externalizing behaviors (e.g. rule-breaking, aggression and conduct problems), internalizing behaviors (e.g. suicidality and youth reported depression), psychosis-related behaviors (e.g. prodromal psychosis, bipolar symptoms and thought problems), and inattentive and social problems. Given the correlation between behavioral problems in youth, this supports evidence that these frequently comorbid behaviors across different behavioral domains have shared genetic influences^5,40,41^. This indicates a common pathway that may contribute to the development of psychopathology. Indeed, suicidality and depression are common across individuals with several different psychiatric disorders and there is evidence that externalizing behaviors in childhood may indicate risk for both externalizing and internalizing disorders in adulthood^42,43^. However, there was some specificity in the behaviors predicted by the ADHD and DEP PRS. The ADHD PRS specifically associated with behavioral approach subscales, impulsivity, ADHD symptoms and mania; whereas the DEP PRS associated with somatic complaints, insomnia, anxiety symptoms, impulsivity specifically in response to negative affect and behavioral inhibition. When controlling for other measures of genetic risk (FH and PRS) in the same model, the PRS-behavior associations became more specific. These findings highlight potentially distinct pathways associated with the development of these unique profiles of behaviors.

Our results replicated previous findings, with a similar magnitude of effects, showing that ADHD PRS significantly associated with hyperactive and inattentive traits in a developmental sample^44–47^. Across the PRS, ADHD and ASD were moderately correlated, and when controlling for the other genetic predictors ASD no longer associated with behavioral problems on the CBCL highlighting the genetic overlap between these disorders in development^48^. There may be additional factors that contributed to the lack of unique relationship of ASD PRS to youth behaviors. Not attending mainstream school classes and an inability to carry out the ABCD protocol, which includes a two-hour MRI scan, was an exclusion criterion; therefore, many individuals with low functioning ASD would have been ineligible for the study. This suggests that the prevalence of ASD symptoms in the ABCD cohort is likely small and restricted to only part of the autism spectrum, which may have a larger overlap with ADHD. Moreover, only 17% of the phenotypic variance in ASD is thought to be attributable to common genetic variants at current sample sizes^49^ and our PRS was not sensitive to important rare chromosome deletions in ASD^50^.

Interestingly, in our sample, the ADHD PRS predicted many bipolar-related behaviors and psychotic-like symptoms. Symptom profiles for pediatric BPD and ADHD are very similar and there is high comorbidity across these disorders^51^. Other studies have shown that childhood ADHD is often a premorbidity for the later development of schizophrenia and relatives of individuals with schizophrenia have higher rates of ADHD than the general population^52–54^. Given the low correlation between ADHD, SCZ and BPD PRS in this study, the ADHD PRS may highlight individuals at risk for developing psychosis-related disorders that may be etiologically distinct from those with high SCZ or BPD scores.

Despite previous studies showing that the SCZ PRS associates with several markers of general psychopathology in adolescence^8,45,55,56^, we did not find any associations of SCZ or BPD PRS with psychopathology in our models. This could be driven by differences in the statistical approach, the demographics of the samples or the phenotypes measured – which can impact the stability of results across adolescent samples^57^. Nevertheless, as hypothesized, we did identify a significant negative association between the SCZ PRS and the fluid composite score from the NIH Toolbox®, which remained after controlling for sociodemographic factors and was unique to the SCZ PRS. Cognitive impairment is a core feature of several psychiatric disorders, particularly those that include psychotic symptoms. Neurodevelopmental studies have highlighted premorbid cognitive impairment across domains in patients with schizophrenia and bipolar disorder^58,59^. Indeed, there is a large genetic overlap across schizophrenia, bipolar disorders and general intelligence^9,10^, which suggests there are shared etiological mechanisms that affect psychopathology and cognition. Perturbations in the cognitive domain may indicate early risk for schizophrenia more readily than other behavioral manifestations.

There were differences in associations across caregiver and youth reported behaviors, particularly with genetic risk for depression. In the multivariable models, youth-reported depression symptom scores were more associated with the DEP PRS, whilst caregiver-reported depression was associated with a FH of depression. Informant discrepancies between caregiver and child-reported measures have been widely reported^60^ and we see relatively low correlations between youth and caregiver reported measures in the current study. Negative biases from caregivers, particularly due to caregiver depression, can also impact behavioral reports^16,61^. An awareness of a history of depression within the youth’s family may have biased the informant’s report about the youth’s depression, leading to a stronger relationship of FH of depression with caregiver compared to youth reported measures. Future time points are required to delineate which informant-reported measures are more accurate at predicting later diagnoses.

FH of anxiety/stress and conduct problems showed the greatest number of associations across different behavioral domains, supporting a role for anxiety or sensitivity to stress and delinquent behavior as transdiagnostic traits. However, in the multivariable models in particular, there were subtle differences in the pattern of FH-behavior associations across domains. For example, FH of drug abuse explained unique variance in rule-breaking behaviors; whereas FH of alcohol abuse explained unique variance in social problems and anxious/depressive behaviors. This highlights that the pattern of behaviors across domains associated with a predictor, when accounting for other measures of genetic risk, is likely important for understanding predictive specificity across disorders. Inherent to FH measures are implicit genetic and environmental influences that are difficult to separate. It remains to be seen whether the additional variance in behavior explained by FH measures above and beyond PRS reflects environmental or additional genetic influences. Together FH and PRS measures predicted ∼6% of the variability in the CBCL Total Problems score. These analyses highlight the utility of measuring multiple markers of genetic risk.

There are several limitations in the current study. PRS association strength is limited by the phenotype’s heritability and the training sample used^62,63^. DEP had the largest discovery sample (Supplementary Figure 1) and the lowest SNP heritability, yet displayed some of the largest associations in our sample. This may be due to depression having relatively greater population prevalence compared to the other psychiatric disorders measured, therefore compared to other disorders, risk alleles may be well represented in our sample. The correlations between the PRS generated in this study were much lower than the genetic correlations determined in the original GWAS, which may be because this cohort is not enriched for individuals with risk alleles. Many psychiatric disorders have increased penetrance during adolescence, therefore the lack of variance in psychopathology symptoms at this age may explain the limited associations between behavior and the SCZ/BPD PRS. Moreover, the GWAS used to produce the PRS in this study were all conducted on European samples. The ABCD sample is demographically diverse, however PRS trained in different ancestry groups do not validly predict phenotypes in admixed or different ancestry samples. This highlights the limited predictive capacity of European-only GWAS for admixed populations and emphasizes the need for conducting GWAS in different ancestry groups. Finally, the magnitude of the genetic-behavior effects detected was very small; the development of psychopathology is extremely complex and genetic risk as estimated with polygenic predictors appears to only account for a small proportion of variability in behavior at this age.

Here we have shown that different PRS and FH measures predicted unique patterns of symptoms of psychopathology, related individual difference factors and cognitive function in a large sample of 9 to 11-year-old children. The unique associations controlling for other genetic measures provides encouraging evidence that genetic data may be useful alongside FH in identifying specific risk for psychiatric disorders. Longitudinal analyses will further elucidate the specificity of these associations and can track these coexisting patterns of behavior to determine the differential predictive utility for PRS and FH measures.

## Data Availability

All data are available to download from the NIMH Data Archive ABCD Collection Release 2.0.1 (DOI: 10.15154/1504041).

## ACKNOWLEDGEMENTS

The authors wish to thank the youth and families participating in the Adolescent Brain Cognitive Development (ABCD) Study and all ABCD staff. Data used in the preparation of this article were obtained from the Adolescent Brain Cognitive Development (ABCD) Study (https://abcdstudy.org), held in the NIMH Data Archive (NDA). This is a multisite, longitudinal study designed to recruit more than 10,000 children age 9-10 and follow them over 10 years into early adulthood. The ABCD Study is supported by the National Institutes of Health and additional federal partners under award numbers U01DA041022, U01DA041028, U01DA041048, U01DA041089, U01DA041106, U01DA041117, U01DA041120, U01DA041134, U01DA041148, U01DA041156, U01DA041174, U24DA041123, U24DA041147, U01DA041093, and U01DA041025. A full list of supporters is available at https://abcdstudy.org/federal-partners.html. A listing of participating sites and a complete listing of the study investigators can be found at https://abcdstudy.org/Consortium_Members.pdf. ABCD consortium investigators designed and implemented the study and/or provided data but did not all necessarily participate in analysis or writing of this report. This manuscript reflects the views of the authors and may not reflect the opinions or views of the NIH or ABCD consortium investigators. The ABCD data repository grows and changes over time. The data was downloaded from the NIMH Data Archive ABCD Collection Release 2.0.1 (DOI: 10.15154/1504041). C.F. was supported by grant R01MH122688 funded by the National Institute for Mental Health (NIMH).

