## Supplementary Material for "Unique prediction of developmental psychopathology from genetic and familial risk"

1. Supplementary methods
2. Supplementary Tables 1-4
3. Supplementary Figures 1-8
4. Statistical data tables
5. Supplementary material references

#### Supplementary Methods

##### *Genetic Data Preprocessing*

At the baseline visit blood or saliva samples of participants were collected and sent to Rutgers University Cell and DNA Repository for DNA isolation and storage. After profile checking and DNA sample preparation quality control to ensure sample validity, genotyping was performed on 646,247 genetic variants using the Affymetrix smokescreen array[1]. Successful genotype calls were determined based on the recommendation of Affymetrix Axiom Analysis Suite v5.0, with at least 98% call rates. Further quality controls were implemented after merging of genotyping batches for all ABCD participants, including inbreeding check, sex concordance check, and cohort level missingness check. After batch specific QC and cohort level QC, 1,221 individuals and 128,523 markers were removed.

We derived genetic ancestry factors (GAFs) using fastStructure with four ancestry factors[2] and genetic relatedness was computed using PLINK[3]. Imputation was performed using the Michigan Imputation Server[4] with hrc.r1.1.2016 reference panel, Eagle v2.3 phasing and multiethnic imputation process. Best guess conversion at a threshold of 0.9 was used to convert dosage files to plink files using PLINK[3]. Post imputation QC criteria was an imputation quality score greater than 0.9 and a Hardy-Weinberg threshold of $10^{-6}$. This QC filtering was performed using PLINK[3] and resulted in 1,427,972 remaining markers and 10,659 individuals.

##### *PRS estimation (additional details)*

Due to linkage disequilibrium nearby SNPs are often correlated with one another, as such these are removed before polygenic scoring, this process is known as clumping and pruning. After genetic imputation and post imputation QC, we performed clumping of SNPs using PRSice[5] with a clumping $r^2$ of 0.1 , clumping window of 250 kb. We did not use a p-value threshold for calculating the polygenic scores, except for DEP which only included the top 10k SNPs due to legal stipulations of the 23andMe sample – this induced an effective threshold of ~0.003. Additionally, variants part of the major histone compatibility (MHC) region (chromosome 6 28MB-34MB) were removed from the analysis due to its highly variable LD structure[6]. However, we retained the C4 locus (situated in the MHC region) due to its strong association with schizophrenia[7,8]. Idels and multi-allelic SNPs were excluded. The PRS for each participant was calculated as the dot product of the allele value at each loci multiplied by its effect size. Due to some polygenic scores having skewed distributions we decided to rank normalize[9] each score to ensure they followed a normality.

##### *Statistical Analysis (additional details)*

Due to convergence issues when using mixed effects models with a random effect of family, we controlled for family relatedness using a random subset of the sample that only included singletons. To ensure the stability of our findings we ran all models in 100 random subsamples of singletons and took the median of effect sizes across all iterations. We calculated effect sizes as $R^2$ type III sum of squares using Nagelkerke's correction to Cox and Snell's formulation[10,11]. P-values were calculated using a log likelihood ratio test and significant associations determined using a false discovery rate (FDR) significance threshold calculated using the Benjamini-Hochberg method[12]. This correction was made within each ancestry group and model type (univariate or multivariate) – i.e. correcting for 15 genetic/familial predictors and 41 behavioral assessments = 615 multiple tests. GLMs were implemented using the R stats package.  Model output for all averaged models is downloadable as csv files (see Statistical Data Tables).

The distribution of each of the DVs fell into three categories a) normal, b) right skewed, zero inflated or c) binary, and we appropriately modelled each of these distributions differently. A) For normal distributions we further ensured normality by rank normalizing[9] (as was performed for PRS), and fit GLMs using the default gaussian family. B) For the right skewed, zero inflated distributions we fit using a gamma distribution with a log link function, first ensuring that each distribution was non-negative to ensure correct bounds for the link function. C) For binary variables we performed logistic regression.  KSADS symptom scores (except for the youth and caregiver reported total symptom score) were binarized using a median split, as they exhibited convergence issues when treated as continuous.

***Associations across ancestry strata***

As PRS were trained on European individuals and ABCD has high ancestral admixture, the main analyses were performed in a European only sample (European Genetic Ancestry Factor (EUR-GAF)>0.9) and supplementary analyses were conducted in the full sample and a non-European sample (EUR-GAF<0.9) to check for consistency across ancestral groups.  Allele frequency differences across ancestral groups can lead to spurious results when PRS trained on a single ancestral group are applied to samples of different or mixed genetic ancestry. Supplementary Figure 6 shows the same univariate and multivariable PRS and FH associations in the full sample (n=9,168 with complete genetic data) and a non-European sample (n=3,964).  There was a similar pattern of associations for the full sample compared to the Europeans, but with far fewer significant associations for the non-European sample, despite very similar prevalence rates across KSADS diagnoses for the three samples (Supplementary Table 1 and 2). Supplementary Figure 7 shows broadly consistent analogous associations between European and non-European groups; however, there was moderate dispersion observed between estimated effect sizes between groups. These effects are difficult to interpret as the discovery sample only included European individuals. These inconsistencies once again demonstrate the issues of portability of GWAS between ancestry groups[13–15].

| Questionnaire | Variables Analyzed | DEAP Variable Names | Informant | Domain |
|---|---|---|---|---|
| Child Behavior Checklist (CBCL) | CBCL Aggressive<br>CBCL Anxious/Depressive<br>CBCL Rule-breaking<br>CBCL Inattention<br>CBCL Social Problems<br>CBCL Thought Problems<br>CBCL Somatic Complaints<br>CBCL Withdrawn/Depressive<br>CBCL Total Problems | cbcl_scr_syn_aggressive_r<br>cbcl_scr_syn_anxdep_r<br>cbcl_scr_syn_rulebreak_r<br>cbcl_scr_syn_attention_r<br>cbcl_scr_syn_social_r<br>cbcl_scr_syn_thought_r<br>cbcl_scr_syn_somatic_r<br>cbcl_scr_syn_withdep_r<br>cbcl_scr_syn_totprob_r (sum of all sub-scales) | Caregiver | Externalizing<br>Internalizing<br>Externalizing<br>Externalizing<br>Developmental<br>Psychosis-related<br>Internalizing<br>Internalizing<br>NA |
| General Behavior Inventory | General Behavior Inventory - Mania | pgbi_ss_score_p | Caregiver | Psychosis-related |
| Prosocial Behavior Survey (youth) | Prosociality | prosocial_ss_mean | Youth | Prosociality |
| Prodromal Questionnaire Brief Version (PQ-B) | Prodromal Pyschosis Severity Score | prodrom_psych_ss_severity_score | Youth | Psychosis-related |
| UPPS-P for Children Short Scale | UPPS Lack of Perseverance<br>UPPS Lack of Planning<br>UPPS Positive Urgency<br>UPPS Negative Urgency<br>UPPS Sensation Seeking | upps_ss_lack_of_perseverance<br>upps_ss_lack_of_planning<br>upps_ss_positive_urgency<br>upps_ss_negative_urgency<br>upps_ss_sensation_seeking | Youth | Externalizing<br>Externalizing<br>Externalizing<br>Externalizing<br>Externalizing |
| Behavioral inhibition and behavioral activation (BISBAS) scale | BISBAS Drive<br>BISBAS Fun Seeking<br>BISBAS Reward Responsiveness<br>BISBAS Inhibition | bisbas_ss_bas_drive<br>bisbas_ss_bas_fs<br>bisbas_ss_bas_rr<br>bisbas_ss_bis_sum | Youth | Externalizing<br>Externalizing<br>Externalizing<br>Internalizing |
| Kiddie Schedule for Affective Disorders and Schizophrenia (KSADS) categorical diagnostic assessments | KSADS Symptoms Depression<br>KSADS Symptoms Bipolar<br>KSADS Symptoms Anxiety<br>KSADS Symptoms OCD<br>KSADS Symptoms Eating Disorder<br>KSADS Symptoms ADHD<br>KSADS Symptoms Oppositional/Conduct<br>KSADS Symptoms Developmental Disorders<br>KSADS Symptoms PTSD<br>KSADS Symptoms Insomnia<br>KSADS Symptoms Suicidality<br>KSADS Total Symptoms | Modules 1 (depressive disorder) & 3 (disruptive mood dysregulation)<br>Module 2 (all bipolar subtypes)<br>Module 8 (social anxiety) & 10 (general anxiety)<br>Module 11 (OCD)<br>Module 13 (eating disorders)<br>Module 14 (ADHD)<br>Module 15 (oppositional defiant) & 16 (conduct disorder)<br>Module 18 (other developmental disorder NOT autism)<br>Module 21 (PTSD)<br>Module 22 (sleep problems)<br>Module 23 (suicidality)<br>Summary score across all modules | Caregiver & Youth<br>Caregiver & Youth<br>Caregiver & Youth<br>Caregiver<br>Caregiver<br>Caregiver<br>Caregiver<br>Caregiver<br>Caregiver<br>Caregiver & Youth<br>Caregiver & Youth<br>Caregiver & Youth | Internalizing<br>Psychosis-related<br>Internalizing<br>Internalizing<br>Internalizing<br>Externalizing<br>Externalizing<br>Developmental<br>Internalizing<br>Sleep<br>Internalizing<br>NA |
| NIH Toolbox® | NIH Toolbox® Fluid Composite Score<br>NIH Toolbox® Crystallized Composite Score | nihtbx_cryst_uncorrected<br>nihtbx_fluidcomp_uncorrected | Youth<br>Youth | Cognition<br>Cognition |

***Supplementary Table 1.*** *DEAP variable names for all behavioral variables analysed in this study. The KSADS symptom scores were calculated by summing all of the symptom items in each KSADS module indicated in the table. Symptom items were all binary (1 or 0). Both past and present symptoms were included in the summary scores, therefore summary scores represent a lifetime assessment of symptoms associated with a particular disorder. Symptom summary scores were calculated by summing the symptoms within each module (the exception was for ODD and CD which were summed together). All KSADS symptom scores were then coded as binary using a median split for statistical modelling. Behavioral domain represents broad categories of construct similarity used for data visualization. R code to extract and process the variables used for this analysis will be published on the ABCD study GitHub: https://github.com/ABCD-STUDY/.*

| Behavior | DEAP Variable Name Lead Q | Lead Question |
|---|---|---|
| Alcohol use | **famhx_4_p** | Has ANY blood relative of your child ever had any problems due to alcohol, such as: Marital separation or divorce; Laid off or fired from work; Arrests or DUIs; Alcohol harmed their health; In an alcohol treatment program; Suspended or expelled from school 2 or more times; Isolated self from family, caused arguments or were drunk a lot. |
| Drug use | **fam_history_5_yes_no** | Has ANY blood relative of your child ever had any problems due to drugs, such as: Marital separation or divorce; Laid off or fired from work; Arrests or DUIs; Drugs harmed their health; In a drug treatment program; Suspended or expelled from school 2 or more times; Isolated self from family, caused arguments or were high a lot. |
| Depression | **fam_history_6_yes_no** | Has ANY blood relative of your child ever suffered from depression, that is, have they felt so low for a period of at least two weeks that they hardly ate or slept or couldn't work or do whatever they usually do? |
| Mania | **famhx_7_yes_no** | Has ANY blood relative of your child ever had a period of time when others were concerned because they suddenly became more active day and night and seemed not to need any sleep and talked much more than usual for them? |
| Psychosis (visions) | **famhx_8_yes_no** | Has ANY blood relative of your child ever had a period lasting six months when they saw visions or heard voices or thought people were spying on them or plotting against them? |
| Conduct problems (trouble) | **famhx_9_yes_no** | Has ANY blood relative of your child been the kind of person who never holds a job for long, or gets into fights, or gets into trouble with the police from time to time, or had any trouble with the law as a child or an adult? |
| Nerves | **famhx_10_yes_no** | Has ANY blood relative of your child ever had any other problems with their nerves, or had a nervous breakdown? |
| Seen a therapist (professional) | **famhx_11_yes_no** | Has ANY blood relative of your child ever been to a doctor or a counselor about any emotional or mental problems, or problems with alcohol or drugs? |
| Hospitalized | **famhx_12_yes_no** | Has ANY blood relative of your child ever been hospitalized because of emotional or mental problems, or drug or alcohol problems? |
| Suicide | **famhx_13_yes_no** | Has ANY blood relative of your child ever attempted or committed suicide? |

***Supplementary Table 2.*** *Description of the family history variables used and the questions asked. If the participant had ANY blood relative who endorsed the described behavior the "DEAP variable name Lead Q" was endorsed with a 1 (if not = 0).*

| | PRS Effect (beyond FH + covariates) | | | FH Effect (beyond PRS + covariates) | | | PRS + FH Effect (beyond covariates) | | |
|---|---|---|---|---|---|---|---|---|---|
| DV | $\Delta R^2$ | $\chi^2$ | $p(\chi^2)$ | $\Delta R^2$ | $\chi^2$ | $p(\chi^2)$ | $\Delta R^2$ | $\chi^2$ | $p(\chi^2)$ |
| **BISBAS Drive (Y)** | 0.14% | 4.92 | 1.08E-01 | 0.20% | 7.21 | 2.11E-01 | 0.35% | 12.37 | 9.07E-02 |
| **BISBAS Fun Seeking (Y)** | 0.16% | 7.64 | 2.22E-01 | 0.26% | 12.25 | 3.42E-01 | 0.43% | 20.63 | 2.20E-01 |
| **BISBAS Reward Responsiveness (Y)** | 0.37% | 33.05 | 6.39E-03 | 0.12% | 10.33 | 8.88E-01 | 0.49% | 43.81 | 1.24E-01 |
| **BISBAS Inhibition (Y)** | 0.25% | 10.37 | 5.57E-02 | 0.41% | 17.08 | 5.87E-02 | 0.66% | 28.04 | 1.52E-02 |
| **UPPS Lack of Perseverance (Y)** | 0.17% | 6.07 | 3.05E-02 | 0.34% | 12.18 | 5.86E-03 | 0.54% | 19.33 | 5.85E-04 |
| **UPPS Lack of Planning (Y)** | 0.35% | 13.74 | 9.14E-03 | 0.30% | 11.78 | 2.17E-01 | 0.68% | 26.66 | 1.31E-02 |
| **UPPS Positive Urgency (Y)** | 0.10% | 4.31 | 1.61E-01 | 0.33% | 14.73 | 2.55E-03 | 0.46% | 20.35 | 1.11E-03 |
| **UPPS Negative Urgency (Y)** | 0.30% | 11.98 | 2.35E-02 | 0.27% | 10.91 | 2.96E-01 | 0.60% | 24.25 | 3.50E-02 |
| **UPPS Sensation Seeking (Y)** | 0.24% | 10.92 | 6.17E-02 | 0.26% | 11.79 | 3.30E-01 | 0.49% | 22.31 | 1.22E-01 |
| **CBCL Total Problems (C)** | 0.61% | 21.00 | 6.50E-05 | 5.27% | 190.82 | 1.86E-46 | 6.17% | 225.69 | 2.73E-52 |
| **CBCL Aggressive (C)** | 0.32% | 22.45 | 1.12E-02 | 2.34% | 169.31 | 2.51E-19 | 2.81% | 204.09 | 2.72E-21 |
| **CBCL Anxious/Depressive (C)** | 0.18% | 11.42 | 9.41E-02 | 3.12% | 197.88 | 8.37E-30 | 3.43% | 218.15 | 2.96E-30 |
| **CBCL Rule Breaking (C)** | 0.69% | 47.10 | 3.10E-04 | 2.66% | 185.05 | 3.21E-15 | 3.73% | 261.84 | 3.43E-20 |
| **CBCL Inattention (C)** | 0.65% | 44.36 | 2.05E-06 | 2.03% | 140.32 | 1.02E-18 | 2.85% | 198.87 | 4.03E-25 |
| **CBCL Social Problems (C)** | 0.26% | 18.12 | 9.82E-02 | 2.62% | 189.85 | 1.90E-16 | 3.04% | 220.84 | 3.97E-17 |
| **CBCL Thought Problems (C)** | 0.26% | 15.50 | 3.54E-02 | 3.06% | 188.48 | 3.28E-26 | 3.43% | 212.51 | 3.99E-27 |
| **CBCL Somatic Complaints (C)** | 0.42% | 26.50 | 1.25E-03 | 2.65% | 171.39 | 6.51E-23 | 3.17% | 205.64 | 2.30E-25 |
| **CBCL Withdrawn/Depressive (C)** | 0.23% | 16.31 | 1.84E-01 | 2.31% | 168.37 | 1.43E-12 | 2.59% | 189.32 | 3.09E-12 |
| **Psychosis Severity Score (Y)** | 0.59% | 61.65 | 1.25E-03 | 1.00% | 104.03 | 2.04E-04 | 1.72% | 180.49 | 4.47E-07 |
| **Mania (C)** | 0.23% | 18.83 | 4.06E-01 | 2.99% | 250.72 | 1.24E-10 | 3.41% | 286.29 | 2.19E-10 |
| **Prosociality (Y)** | 0.20% | 6.24 | 1.33E-01 | 0.20% | 6.42 | 5.60E-01 | 0.42% | 13.10 | 2.75E-01 |
| **KSADS Symptoms Depression (C)** | 0.09% | 4.11 | 5.33E-01 | 1.99% | 88.73 | 9.55E-15 | 2.15% | 95.81 | 8.07E-14 |
| **KSADS Symptoms Bipolar (C)** | 0.36% | 16.14 | 6.47E-03 | 1.39% | 61.88 | 1.60E-09 | 1.86% | 82.91 | 2.05E-11 |
| **KSADS Symptoms Anxiety (C)** | 0.30% | 13.43 | 1.97E-02 | 2.35% | 105.28 | 4.76E-18 | 2.64% | 118.11 | 4.39E-18 |
| **KSADS Symptoms OCD (C)** | 0.06% | 2.43 | 7.87E-01 | 1.47% | 65.43 | 3.35E-10 | 1.52% | 67.70 | 1.14E-08 |
| **KSADS Symptoms Eating Disorder (C)** | 0.04% | 1.87 | 8.66E-01 | 0.42% | 18.51 | 4.69E-02 | 0.46% | 20.33 | 1.60E-01 |
| **KSADS Symptoms ADHD (C)** | 0.28% | 12.26 | 3.14E-02 | 1.92% | 85.45 | 4.26E-14 | 2.29% | 102.48 | 4.42E-15 |

| | | | | | | | | | |
|---|---|---|---|---|---|---|---|---|---|
| **KSADS Symptoms Oppositional Conduct (C)** | 0.71% | 31.27 | 8.28E-06 | 1.92% | 85.52 | 4.13E-14 | 2.80% | 125.43 | 1.66E-19 |
| **KSADS Symptoms Developmental Disorders (C)** | 0.12% | 5.30 | 3.81E-01 | 1.55% | 68.84 | 7.43E-11 | 1.67% | 74.51 | 6.93E-10 |
| **KSADS Symptoms PTSD (C)** | 0.09% | 4.00 | 5.50E-01 | 2.09% | 93.50 | 1.08E-15 | 2.27% | 101.22 | 7.65E-15 |
| **KSADS Symptoms Insomnia (C)** | 0.15% | 6.83 | 2.34E-01 | 0.69% | 30.75 | 6.46E-04 | 0.87% | 38.57 | 7.43E-04 |
| **KSADSS Symptoms Suicidality (C)** | 0.35% | 15.39 | 8.82E-03 | 0.98% | 43.54 | 3.98E-06 | 1.41% | 62.75 | 8.45E-08 |
| **KSADS Total Symptoms (C)** | 0.41% | 27.68 | 2.67E-03 | 5.22% | 365.99 | 4.06E-46 | 5.80% | 408.99 | 1.23E-48 |
| **KSADS Symptoms Depression (Y)** | 0.40% | 17.66 | 3.41E-03 | 0.30% | 13.47 | 1.99E-01 | 0.77% | 34.24 | 3.15E-03 |
| **KSADS Symptoms Biploar (Y)** | 0.27% | 11.73 | 3.87E-02 | 0.48% | 21.31 | 1.91E-02 | 0.79% | 34.87 | 2.57E-03 |
| **KSADS Symptoms Anxiety (Y)** | 0.02% | 0.71 | 9.82E-01 | 0.20% | 8.70 | 5.61E-01 | 0.21% | 9.18 | 8.68E-01 |
| **KSADS Symptoms Insomnia (Y)** | 0.12% | 5.37 | 3.73E-01 | 0.14% | 6.37 | 7.83E-01 | 0.27% | 11.92 | 6.85E-01 |
| **KSADS Symptoms Suicidality (Y)** | 0.24% | 10.48 | 6.27E-02 | 0.38% | 16.83 | 7.83E-02 | 0.67% | 29.71 | 1.30E-02 |
| **KSADS Total Symptoms (Y)** | 0.55% | 59.87 | 9.73E-03 | 0.78% | 84.84 | 1.80E-02 | 1.51% | 166.08 | 2.22E-04 |
| **NIHTBX Fluid Composite Score (Y)** | 0.48% | 18.47 | 7.79E-04 | 0.33% | 12.64 | 1.54E-01 | 0.80% | 30.71 | 2.40E-03 |
| **NIHTBX Crystalized Composite Score (Y)** | 0.43% | 15.14 | 2.03E-03 | 0.66% | 23.14 | 1.32E-03 | 1.11% | 39.32 | 1.74E-05 |

***Supplementary Table 3.*** *Unique and shared variance across behaviors predicted by PRS and FH. Change in $R^2$% is calculated by comparing the $R^2$ from a reduced model with covariates of no interest and genetic predictors of no interest, and a full model including the nested reduced model plus the genetic predictors of interest. The $\chi^2$ statistic and p-value from the likelihood ratio test comparing these models is also displayed. The parenthesis in the DV column indicate youth (Y) or caregiver (C) report*

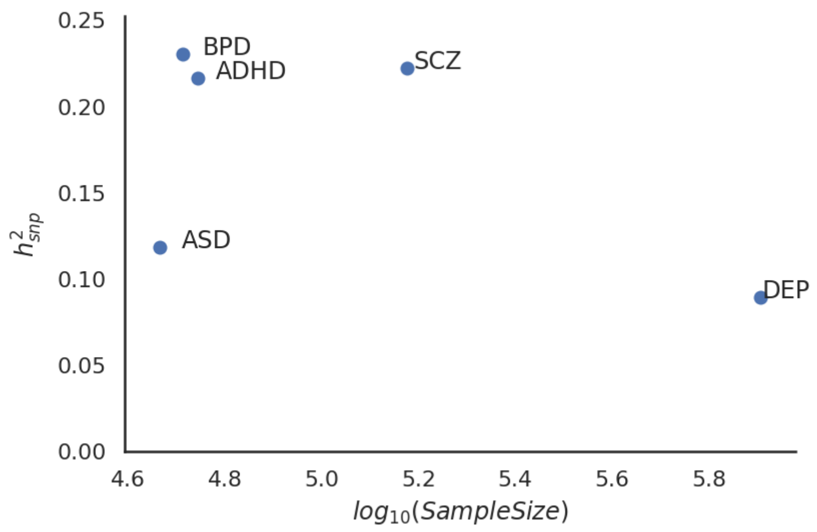


***Supplementary Figure 1.*** *Plot of SNP heritability ($h^2_{snp}$) and log(sample size) for each discovery GWAS sample, as reported in discovery papers, used to calculate PRS.*

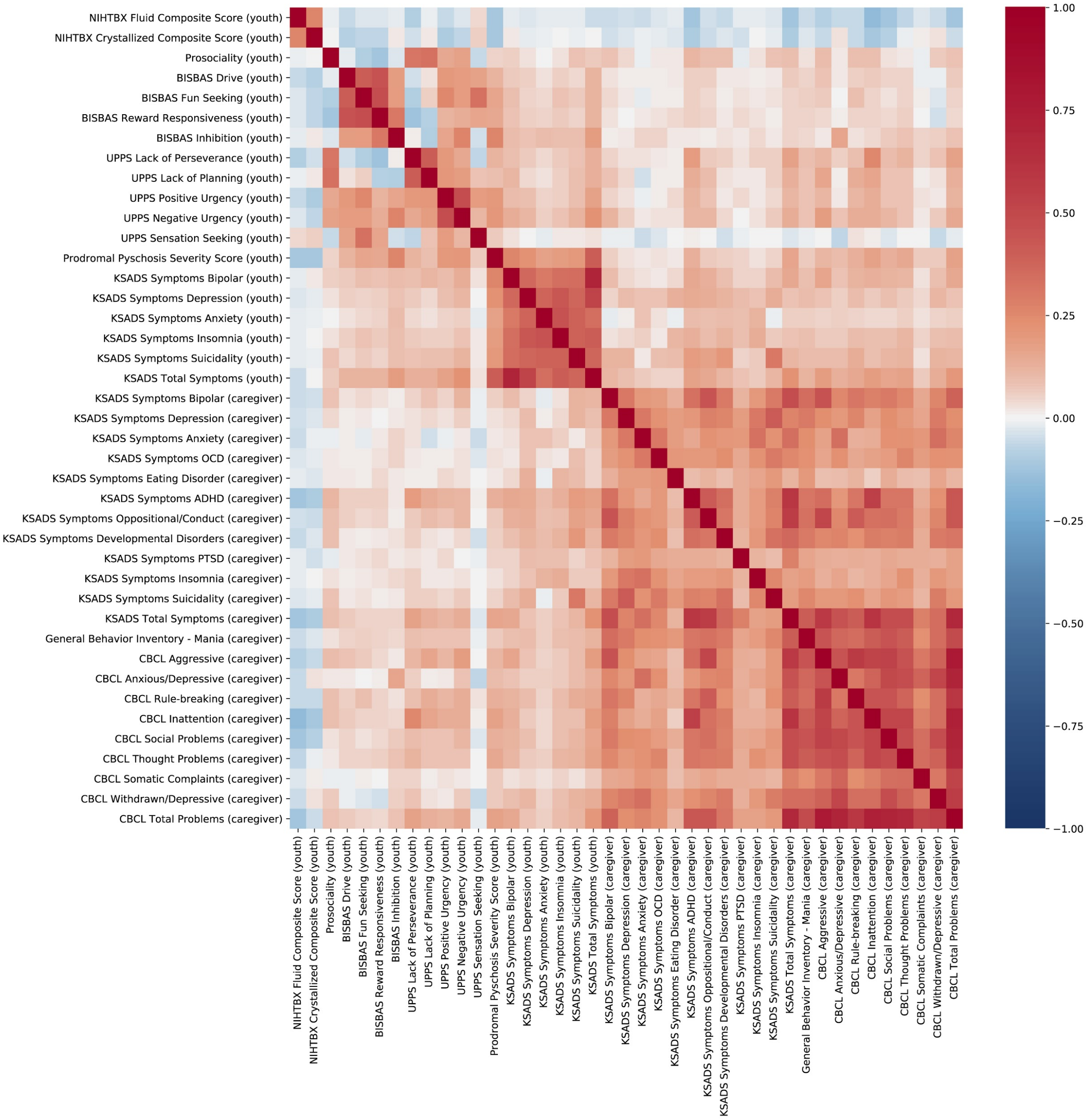


***Supplementary Figure 2.*** *Pairwise spearman correlations between all of the behavioral phenotypes in the European sample pre-residualized for covariates of no interest including socioeconomic status (SES). Caregiver reported measures were more strongly associated with each other than youth reported measures.*

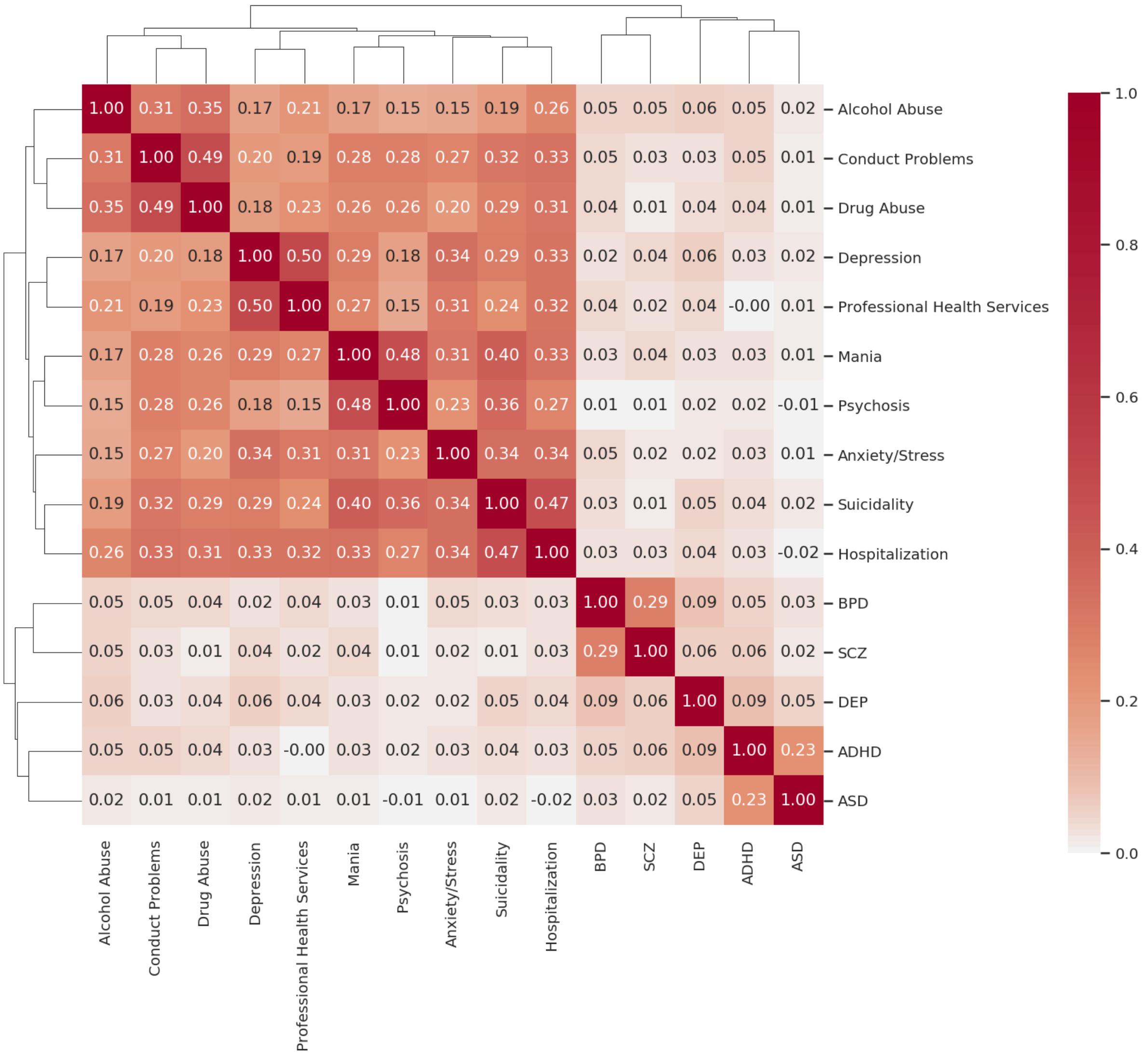


***Supplementary Figure 3.*** *Pairwise spearman correlations between all of the genetic risk measures in the European sample pre-residualized for covariates of no interest including SES. FH measures were more strongly associated with each other than the PRS. The FH measures were moderately correlated with one another. There were limited associations between FH and PRS.*

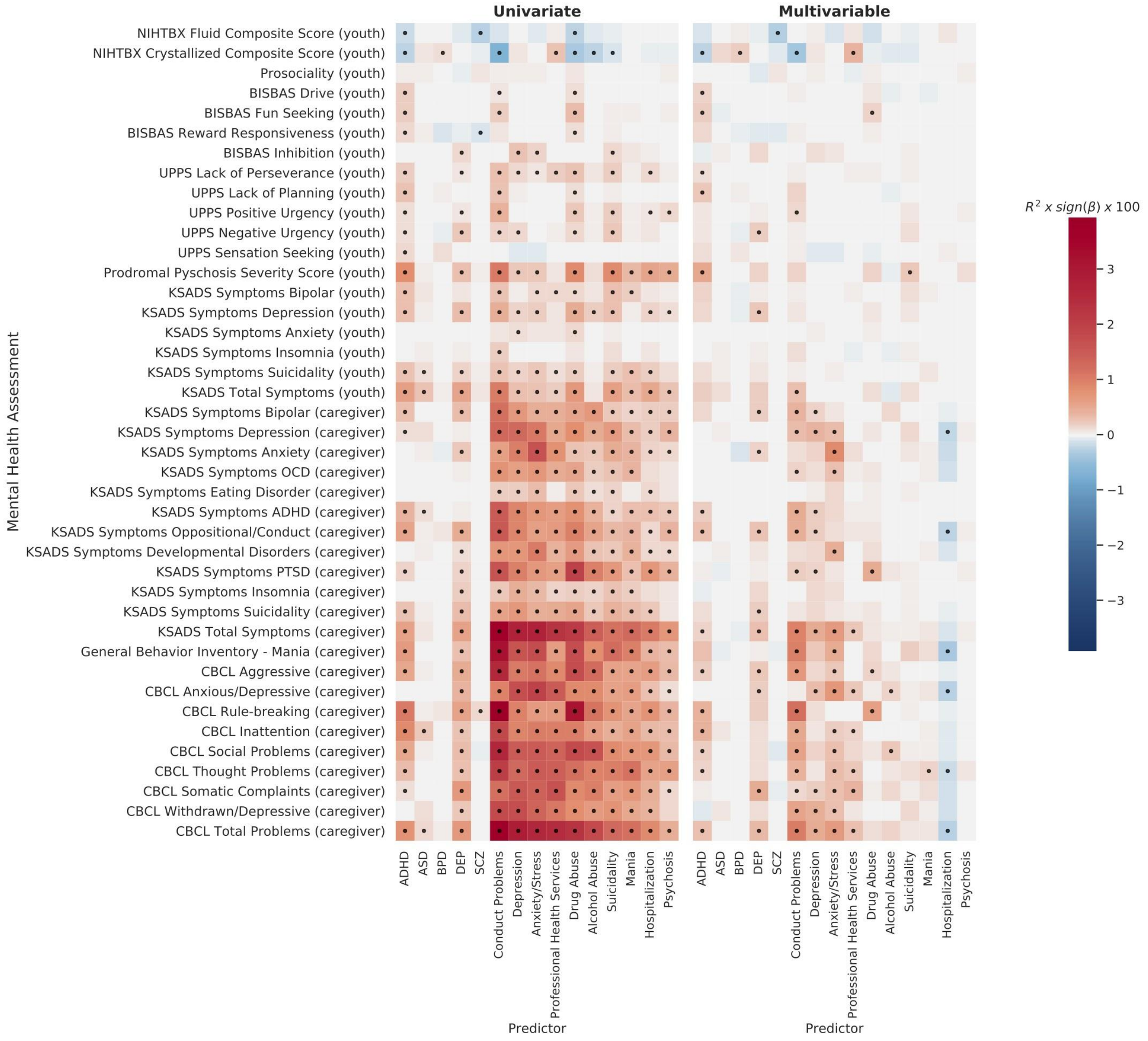


***Supplementary Figure 4. Behavioral associations in the European sample without controlling for SES.*** *Univariate (left) and multivariable (right) associations between the genetic predictors (PRS and Family History) and the behavioral phenotypes, controlling for covariates of no interest, but <u>not</u> controlling for SES. The main differences across the associations with and without controlling for SES were found with the cognitive performance measures. Many associations between genetic risk for psychopathology and cognitive function only reached threshold for statistical significance when SES was not taken into account due to the shared variance between sociodemographic factors and cognition.*

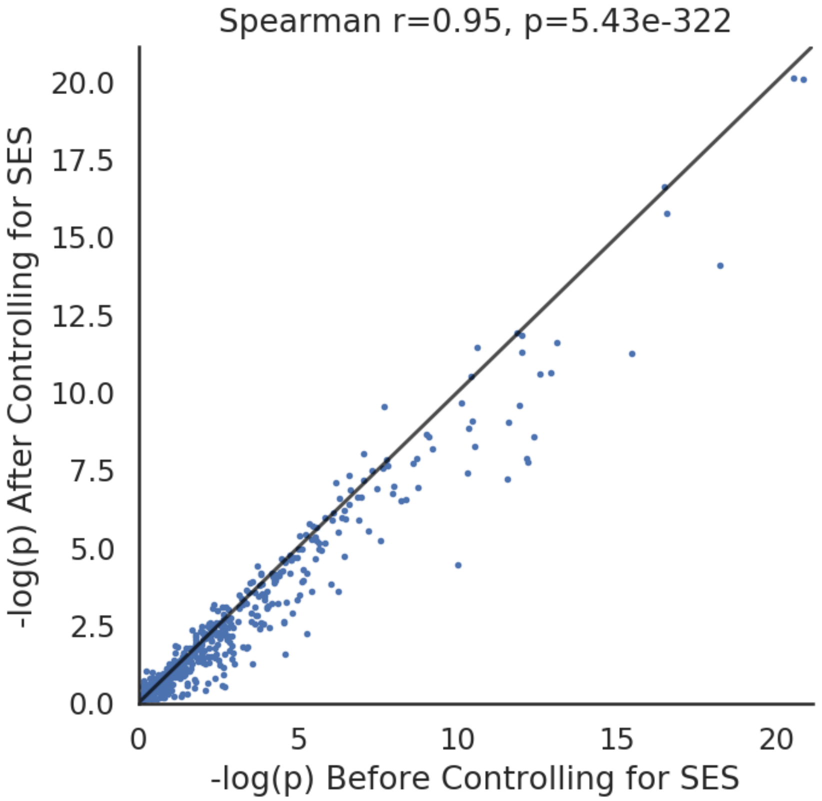


***Supplementary Figure 5.*** *-log(P-values) for all multivariable associations after controlling for SES (Y-axis) and before controlling for SES (X-axis) in the European sample. The pattern of associations was very similar with and without controlling for SES, however controlling for SES attenuated many of the associations.*

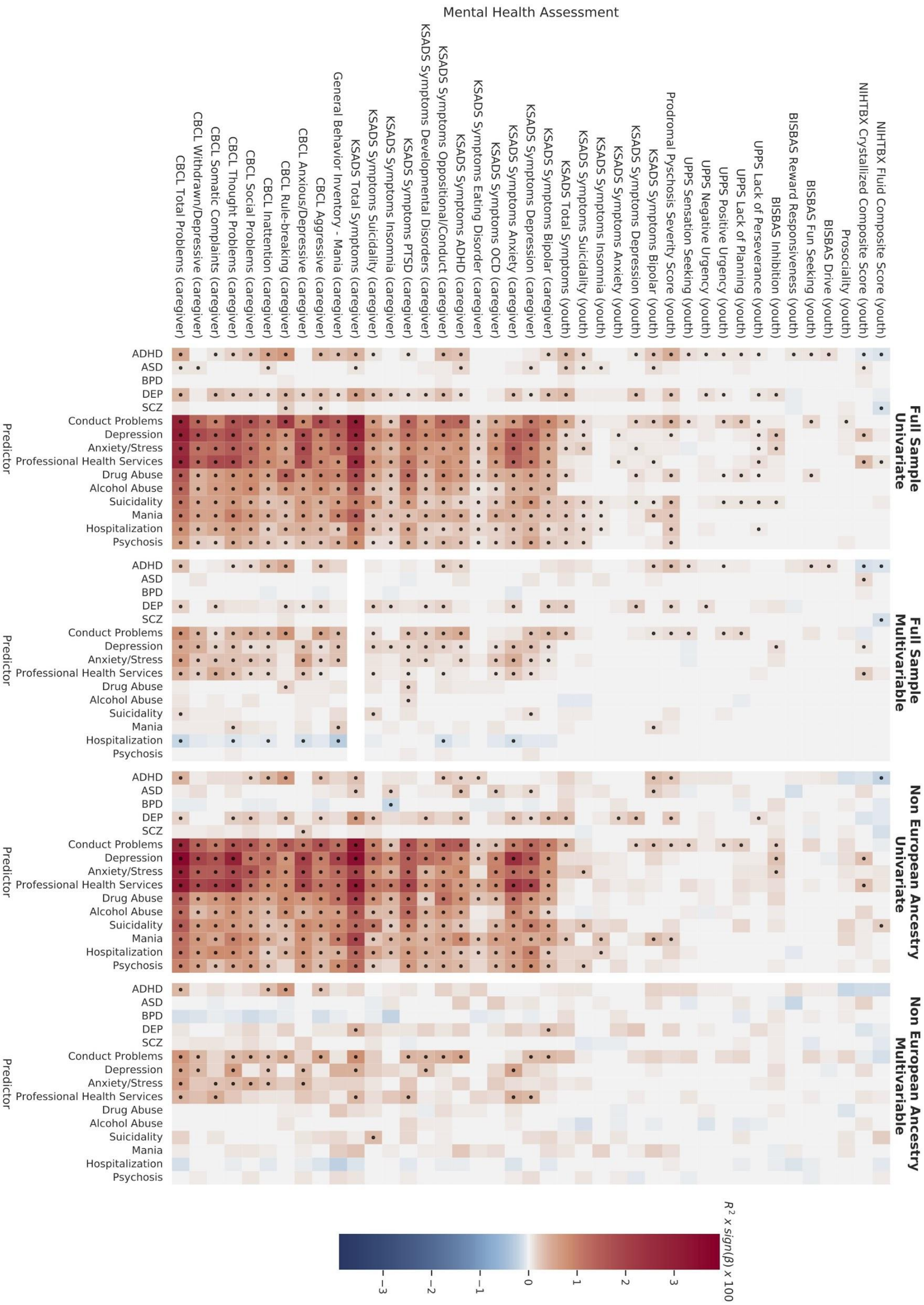


***Supplementary Figure 6. Behavioral associations across ancestry strata.*** *Univariate and multivariable associations between the genetic predictors (PRS and Family History) and the behavioral phenotypes for the full ABCD sample and he non-European ancestry sample, controlling for covariates of no interest including SES. Models that did not converge after 10,000 iterations were left blank.*

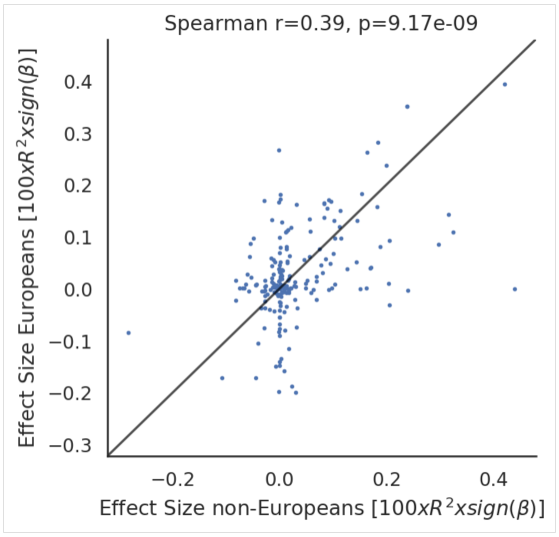


***Supplementary Figure 7.*** *Signed Effect Sizes for all multivariable PRS associations for the European sample (X-axis) and non-European sample (Y-axis) , controlling for covariates of no interest and SES. The estimated associations were broadly consistent between European and non-European groups, however, there was moderate dispersion observed between estimated effect sizes, which highlights difficulties in using PRS generated in certain ancestry groups to other ancestry groups.*

#### Statistical Data Tables

For completeness and transparency of reporting, we have generated additional data tables as .csv files with the results from all statistical models. Each table includes the beta estimate, t statistic, signed percentage variance explained ($R^2$) and p-value for each predictor in each model. These results are the averaged statistics across 100 samples of singletons as outlined in the Supplementary Methods. The following data tables are included:

- EUR Ancestry Results
- Non EUR Ancestry Results
- Full Sample Results

Each file contains a separate sheet for:

1. Univariate SES covaried
2. Multivariable SES covaried multivariable
3. Univariate (not controlling for SES)
4. Multivariable (not controlling for SES)

### Supplementary Material References
